# Geographic Variation in Diagnostic Performance of Amsel Criteria and Nugent Score for Vaginal Dysbiosis Defined by 16S rRNA Gene Sequencing

**DOI:** 10.64898/2026.05.12.26352230

**Authors:** Meilin Zhu, Andile Mtshali, Gugulethu Mzobe, Nzuzo Magini, Nireshni Mitchev, Anam Khan, Briah Cooley Demidkina, Meena Murthy, Lara Lewis, Jiawu Xu, Jonathan Shih, Joseph Elsherbini, Asavela Kama, Nomfuneko A. Mafunda, Callin Chetty, Laura Vermeren, Jo-Ann S. Passmore, Anna-Ursula Happel, Douglas S. Kwon, Laura Symul, Disebo Potloane, Sinaye Ngcapu, Caroline M. Mitchell

**Affiliations:** Ragon Institute of Mass General, Harvard and MIT, Cambridge Massachusetts, United States of America; Centre for the AIDS Programme of Research in South Africa (CAPRISA), University of KwaZulu-Natal, Durban, South Africa; Vincent Center for Reproductive Biology, Massachusetts General Hospital, Boston, Massachusetts, United States of America; Louvain Institute of Data Analysis and Modeling (LIDAM), Université catholique de Louvain, Louvain-la-Neuve, Belgium; Department of Medical Microbiology, School of Medicine, University of KwaZulu-Natal, Durban, South Africa; Centre for Epidemic Response and Innovation (CERI), School of Data Science and Computational Thinking, Stellenbosch University, Stellenbosch, South Africa; Division of Medical Virology and Biomedical Research Institute (BMRI), Faculty of Medicine and Health Science, Stellenbosch University, Tygerberg, Cape Town, South Africa; Division of Medical Virology, Department of Pathology, University of Cape Town, Cape Town, South Africa; Institute of Infectious Disease and Molecular Medicine (IDM), University of Cape Town, Cape Town, South Africa; Division of Immunology, Department of Pathology, University of Cape Town, Cape Town, South Africa; Harvard Medical School, Boston, Massachusetts, United States of America; Division of Infectious Diseases, Massachusetts General Hospital, Boston, Massachusetts, United States of America; Department of Obstetrics & Gynecology, Massachusetts General Hospital, Boston, Massachusetts, United States of America

## Abstract

Vaginal dysbiosis (VD), characterized by low abundance of vaginal lactobacilli and increased bacterial community diversity, is implicated in multiple adverse reproductive outcomes and is an emerging target for preventive interventions, including live biotherapeutic products (LBPs). The most common clinical presentation of VD is bacterial vaginosis (BV), but at least half of people are asymptomatic. We investigated how two commonly used diagnostic criteria for BV, namely Amsel and Nugent, align with 16s rRNA gene sequencing-defined community state types (CSTs) demonstrating VD. We analyzed screening specimens from a Phase 1b randomized trial of LBP conducted at two sites (CAPRISA, South Africa; MGH, USA), as well as a single follow-up visit from enrolled participants. Using sequencing-based CST as the reference and multinomial mixed-effects logistic models, we evaluated the association of Amsel BV and Nugent BV with CST IV (including subtypes IV-A and IV-B) and tested for site-specific effects. Amsel BV was significantly associated with CST IV-A, and IV-B; however, the strength of association was significantly diminished at CAPRISA compared to MGH, pointing to site-specific assessment differences or underlying biological variation. Nugent BV yielded stronger associations with CST IV-A, and IV-B and showed no evidence of a site-specific interaction, indicating consistent performance across sites. These findings indicate that diagnostic performance for VD varies by framework: Amsel criteria are more susceptible to geographical site effects, whereas Nugent score demonstrates stronger and more site-agnostic associations. For clinical studies targeting VD, Nugent scoring and/or sequencing-based approaches should be prioritized for VD endpoint definition and stratification.

**Summary:** Amsel criteria do a poorer job than Nugent score of identifying a diverse, BV-like vaginal microbial community in a South African cohort compared to a North American cohort.

## Introduction

The vaginal microbiome is a key determinant of reproductive health^1-10^ and a promising target for preventive strategies aimed at reducing spontaneous preterm birth, sexually transmitted infection (STI) acquisition (including HIV), infertility, cervical dysplasia, and recurrent urinary tract infections^11-15^. Non-*Lactobacillus*-dominant vaginal communities, hereafter referred to as vaginal dysbiosis (VD), are consistently associated with these adverse reproductive health outcomes, whereas dominance by *Lactobacillus crispatus* is linked to more favorable health states. In contrast, communities dominated by *Lactobacillus iners* show less optimal associations and may represent a less protective or transitional microbiome state^16-18^. As approaches for characterizing the vaginal microbiome technologically evolve, clinicians and researchers must consider which diagnostic frameworks are used in interventional studies targeting VD. Ideally, such criteria should sensitively detect VD while also supporting efficient identification and enrollment of target populations.

The most direct clinical manifestation of VD, bacterial vaginosis (BV), is one of the most common causes of vaginal symptoms such as discharge, irritation, and/or odor^19,20^. The gold standard clinical diagnostic criteria for BV (Amsel criteria) were developed on data from a small US cohort and comprise four findings (elevated vaginal pH, presence of clue cells, positive whiff test, and characteristic discharge), each reflecting functional consequences of an altered vaginal microbiome^21^. Epidemiologic studies more often use microscopy-based Nugent criteria, which score bacterial morphotypes on Gram-stained vaginal smears, and were originally developed to be a reproducible measure of microbiome composition in a US multi-site study of pregnant women^22^. The more recent advances in 16S rRNA gene amplicon sequencing in the context of the vaginal microbiome allows higher resolution identification of bacterial genera and species^23^, enabling assessment of more taxa-specific health relationships, such as the recognition that *L. iners* may not have the same protective associations as other lactobacilli^16-18^. The expanded characterization of the vaginal microbiome enabled by sequencing technologies demonstrates that VD extends beyond the traditional diagnosis of BV, encompassing a spectrum of microbial community states with potential health implications.

Importantly, the non-*Lactobacillus* dominant vaginal microbiomes described by both Nugent scoring and molecular sequencing have been associated with less optimal health outcomes, independent of symptomatic presentation; and these associations have been observed across diverse geographic sites and populations^1-10^. While Amsel criteria define a clinical syndrome, Nugent score and sequencing primarily describe patterns in the vaginal bacterial community. Frequently, the diagnoses returned by these different methods are concordant, but they do not universally agree^24-26^. Additionally, since Amsel’s criteria are used in clinical settings, they are often used only when patients present with symptoms. Studies using Nugent score to screen general populations show that half of people with Nugent BV are asymptomatic, and would, thus, be unlikely to present for care^20,27^. Notably, current diagnostic criteria for Amsel have been developed and validated primarily in Western populations^21,22^. There remains an unmet need to evaluate the applicability and concordance of these criteria for identification of VD across diverse geographical and clinical settings.

To address these gaps, we set out to compare how Nugent score and Amsel criteria relate to the underlying vaginal microbiome composition and assess their reliability in identifying VD in samples collected from two geographically distinct sites – CAPRISA (Centre for the AIDS Programme of Research in South Africa) in Vulindlela, Kwa-Zulu Natal, South Africa and Massachusetts General Hospital (MGH) in Boston, Massachusetts, USA. These samples were collected from VIBRANT (Vaginal lIve Biotherapeutic RANdomized Trial), a Phase 1b clinical trial of a vaginal live biotherapeutic product (LBP)^12^,^28^. To our knowledge, few studies have integrated clinical, microscopic, and molecular diagnostic frameworks to evaluate BV and VD across geographically distinct populations. This work addresses an important gap in the literature and informs the selection of diagnostic strategies for future trials and interventions targeting VD.

## Results

The primary objective of this sub-analysis was to assess how reliably Amsel criteria and Nugent score identified VD and to determine whether these relationships varied by site. VD was defined as CST IV-A and CST IV-B; Amsel BV was defined as having at least 3 positive Amsel criteria components; and Nugent BV was defined as a total Nugent score ≥7. Both sites recruited pre-menopausal women aged 18–40 years.

A total of 532 individuals were screened and 96 were enrolled (26 at MGH and 70 at CAPRISA). Sites differed significantly in the predominant contraceptive method (primarily progestin-only injectable vs. primarily continuous combined oral contraceptive pills; χ^2^ = 332.5, dof = 10, p-value = 2.04E-65; Figure S1A–B). After enrollment, all participants received 7 days of oral metronidazole followed by the investigational product and were followed for 12 weeks. For this analysis, we included all samples from screening and from the week 12 follow-up visit with paired Amsel and Nugent data. A subset also had paired 16S rRNA gene sequencing data. Samples assigned to CST V or IV-C (n=5) were excluded from analyses using 16S rRNA gene sequencing data due to the small number of samples. The total analytic dataset comprised 557 samples (83 from MGH and 474 from CAPRISA) with paired Amsel and Nugent data, representing 474 women (63 from MGH and 411 from CAPRISA), and among these, 228 samples (from 152 individuals) also had available 16S rRNA gene sequencing data (47 samples from 29 individuals at MGH and 181 samples from 123 individuals at CAPRISA). Analyses incorporating sequencing-based CSTs were therefore limited to these 228 samples, whereas analyses based solely on Amsel and Nugent criteria used the full set of 557 samples.

We first examined vaginal microbiome composition in the 228 samples with available sequencing data (Figure 1A). As expected, CST I samples showed low Nugent scores and low Amsel BV positivity, whereas CST III samples exhibited a range of Nugent scores and Amsel positivity. CST IV subtypes were generally characterized by high Nugent scores and high Amsel BV positivity. However, we unexpectedly observed a substantial subset (74/411 [18%]) of CAPRISA samples classified as Nugent BV but Amsel negative.

**Figure 1.**
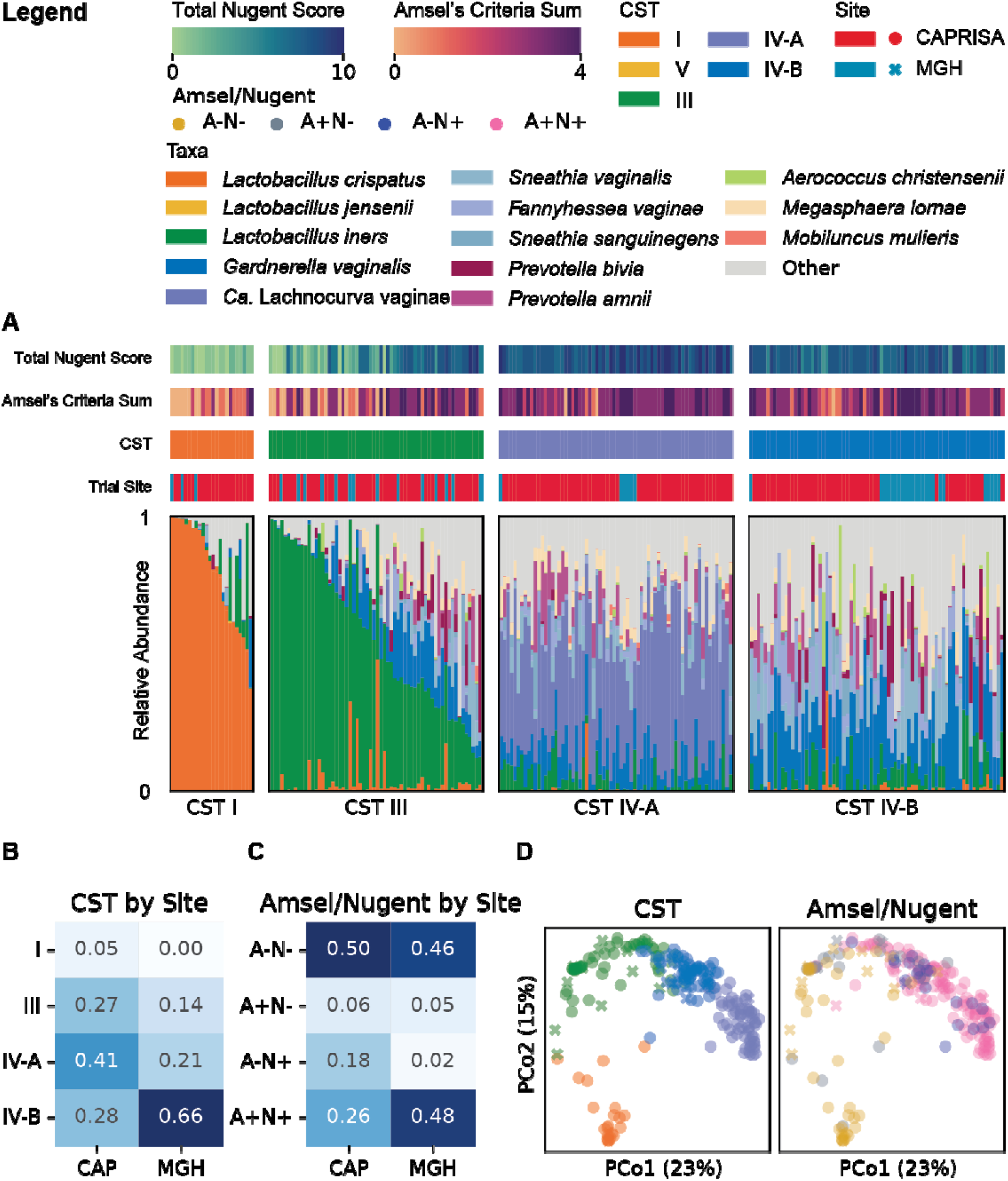
Microbiome composition of screening and study visit samples from VIBRANT across sites. (A) 16s rRNA gene sequencing results of all screening and follow up visit samples included in this analysis from VIBRANT (N=557 total samples; 474 samples from CAP and 83 samples from MGH). (B) Proportion of CST groups per each site for screening samples only (N=152 total samples; 123 samples from CAP and 29 samples from MGH). (C) Proportion of Amsel/Nugent status groups per each site for screening samples only (N=504 total samples; 441 samples from CAP and 63 samples from MGH). (D) PCoA of all samples (N=557), colored by CST (left) or Amsel/Nugent status (right).

When assessing only samples collected from screening visits, CST distributions differed significantly by site (χ^2^=15.2, df=3, p<0.005), with CAPRISA having a higher proportion of CST IV-A samples – characterized by diverse anaerobes with high *Ca*. Lachnocurva vaginae abundance (50/123 [41%] vs. 6/29 [21%], respectively; Figure 1B). We next compared the distribution of diagnostic categories defined by Amsel (A) and Nugent (N) status (A-N-, A+N-, A-N+, and A+N+) across sites. Again, we observed a significant difference between sites (χ^2^=18.5, df=3, p<0.0005), driven largely by a higher proportion of A-N+ samples at CAPRISA compared to MGH (74/411 [18%] vs. 1/63 [2%]; Figure 1C).

To explore whether specific combinations of Amsel and Nugent status corresponded to distinct CST groupings, samples were ordinated using principal coordinate analysis (PCoA) on a Bray-Curtis distance matrix. The first two principal coordinates (PCo1 and PCo2), which together explained 38% of the variation (PCo1: 23%, PCo2: 15%) in the distance matrix, were visualized and colored by Amsel/Nugent status (Figure 1D). A-N-samples overlapped predominantly within CST I and, to a lesser extent, CST III. A+N-samples were rare and did not cluster strongly within any CST. By contrast, A-N+ and A+N+ samples overlapped extensively and spanned the CST IV-A and CST IV-B subregions, with no obvious separation by Amsel BV status within the first 2 principal coordinates. This overlap suggests that the higher prevalence of CST IV-A at CAPRISA alone does not explain the lower proportion of Amsel BV at that site, consistent with the presence of high-Nugent, low-Amsel samples in both CST IV-A and CST IV-B (Figure 1A). Incorporating the third principal coordinate (PCo3, explaining an additional 13% of the variation in the distance matrix) did not materially change these patterns (Figure S2).

To quantitatively compare how Amsel and Nugent criteria relate to CSTs and to test for site-specific effects, we fit Bayesian multinomial mixed-effects logistic regression models to predict CSTs. In one model, the primary predictor was Amsel BV status; in a second model, Nugent BV was evaluated as the predictor. Both Amsel and Nugent BV status were included as binary variables. In both models, BV status (by Amsel or Nugent), site, and a BV status-by-site interaction term were included as fixed effects, and participant was included as a random effect to account for repeated measures of the same individual. CST I served as the reference category for CST, and CAPRISA served as the reference site. We estimated model parameters and corresponding odds ratios (ORs) and credible intervals (CrIs) for each non-reference CST (Tables 1 and 2).

**Table 1.** Model estimates for the Amsel BV model.

| CST | Term | OR | OR lower | OR upper |
| --- | --- | --- | --- | --- |
| CST III | Amsel BV | 1.03E+01 | 2.85E+00 | 4.05E+01 |
| CST III | Site - MGH (ref:CAP) | 6.19E+00 | 1.08E+00 | 5.53E+01 |
| CST III | Amsel BV:Site | 1.40E+03 | 2.95E-02 | 8.79E+12 |
| CST IV-A | Amsel BV | 6.34E+01 | 1.18E+01 | 5.69E+02 |
| CST IV-A | Site - MGH (ref:CAP) | 3.88E-26 | 1.75E-71 | 1.56E-02 |
| CST IV-A | Amsel BV:Site | 1.33E+29 | 2.71E+03 | 1.91E+77 |
| CST IV-B | Amsel BV | 1.21E+01 | 3.46E+00 | 4.87E+01 |
| CST IV-B | Site - MGH (ref:CAP) | 1.08E+00 | 1.29E-01 | 8.71E+00 |
| CST IV-B | Amsel BV:Site | 6.39E+04 | 1.45E+00 | 5.93E+14 |

**Table 2.** Model estimates for the Nugent BV model.

| CST | Term | OR | OR lower | OR upper |
| --- | --- | --- | --- | --- |
| CST III | Nugent BV | 3.58E+02 | 8.84E+00 | 1.10E+06 |
| CST III | Site - MGH (ref:CAP) | 5.09E+00 | 1.05E+00 | 3.52E+01 |
| CST III | Nugent BV:Site | 2.08E+02 | 2.67E-05 | 9.67E+16 |
| CST IV-A | Nugent BV | 8.36E+11 | 1.79E+04 | 2.25E+29 |
| CST IV-A | Site - MGH (ref:CAP) | 1.23E-14 | 1.28E-64 | 8.67E+14 |
| CST IV-A | Nugent BV:Site | 8.36E+16 | 3.63E-14 | 6.06E+70 |
| CST IV-B | Nugent BV | 2.08E+03 | 4.69E+01 | 6.75E+06 |
| CST IV-B | Site - MGH (ref:CAP) | 1.36E+00 | 1.25E-01 | 1.39E+01 |
| CST IV-B | Nugent BV:Site | 1.12E+04 | 1.81E-03 | 8.23E+18 |

In the Amsel-based model, Amsel BV status was significantly associated with CST III (OR 10.3, CrI 2.8–40.5), CST IV-A (OR 63.3, CrI 11.8–569.2), and CST IV-B (OR 12.1, CrI 3.46–48.7). Notably, for CST IV-A and CST IV-B, the strength of this association differed by site, indicating that Amsel BV was less associated with these CST categories in samples from CAPRISA than in samples from MGH (Figure 2A). At CAPRISA, Amsel BV was more common in CST I and CST III samples and less common in CST IV-A and CST IV-B samples (Figure 2B). As expected, based on the CST distributions at each site, MGH site was strongly negatively associated with CST IV-A (OR 3.9×10^-26^, CrI 1.8×10^-71^–0.016).

**Figure 2.**
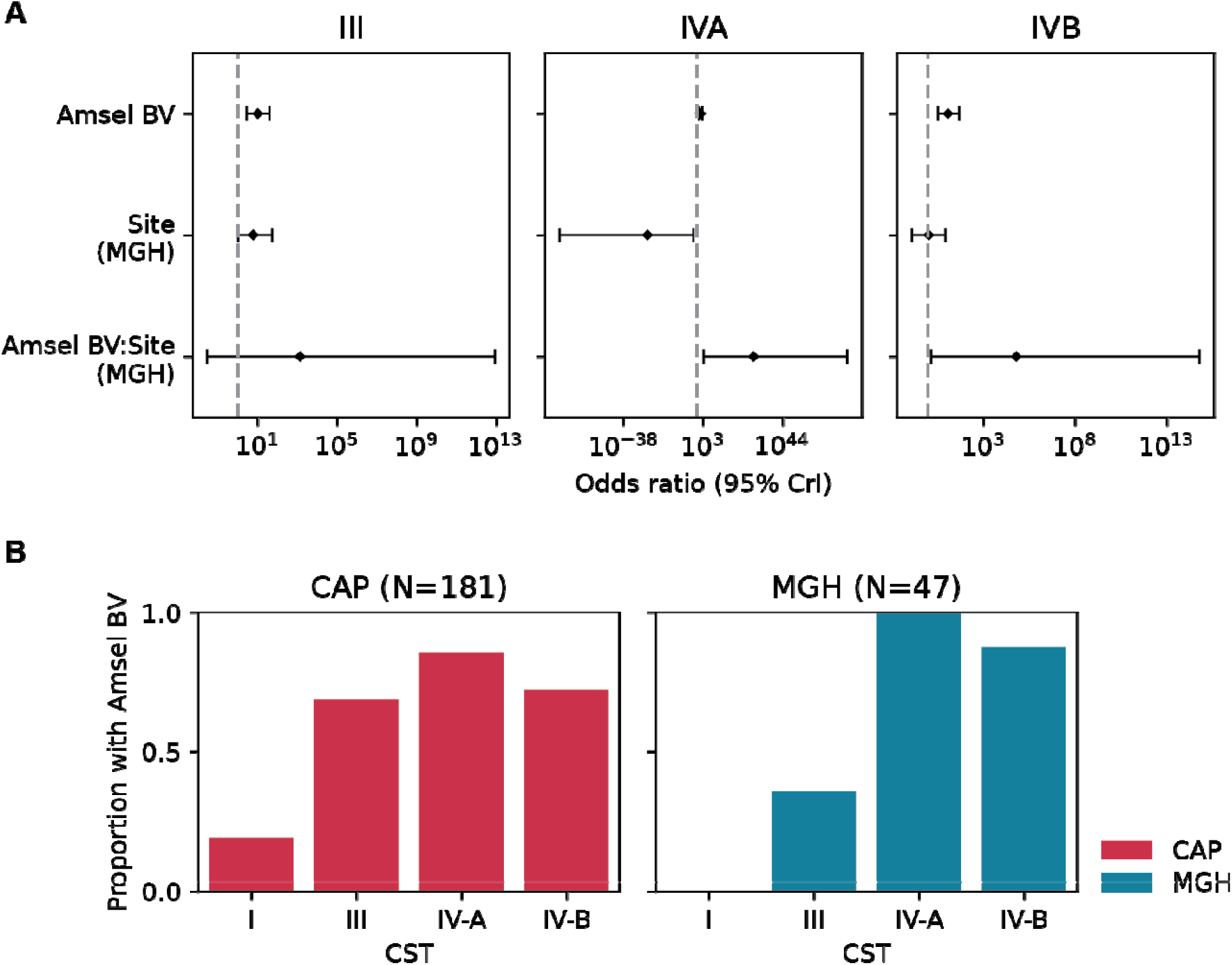
Amsel BV status associates CST with site-specific differences. A) Multinomial odds ratios with 95% credible intervals (CrIs) shown for each predicted CST with CST I as the baseline. Fixed effects included Amsel BV status, the interaction of Amsel BV status and site, and site (with CAPRISA as the baseline). Participant ID was included as a random effect. All screening and follow up visit samples with paired 16s rRNA gene sequencing data were included in this analysis (N=228 total samples; 181 samples from CAP and 47 samples from MGH). B) Proportion of Amsel BV across CST groups for CAPRISA (left) and MGH (right).

In the Nugent-based model, we observed stronger and more consistent associations between Nugent BV and CST membership. Nugent BV was strongly associated with CST III (OR 357.8, CrI 8.8–1.1×10^6^), CST IV-A (OR 8.4×10^11^, CrI 1.8×10^4^–2.3×10^29^), and CST IV-B (OR 2.1×10^3^, CrI 46.8–6.7×10^6^). In contrast to the Amsel-based model, there was no evidence of a site effect on the relationship between Nugent BV and CST III, IV-A, or IV-B, indicating that Nugent scoring identified these CST-defined communities similarly at both sites (Figure 3A). These model-based findings mirrored the observed data, which showed comparable frequencies of Nugent BV among CST IV-A and CST IV-B samples at CAPRISA and MGH (Figure 3B).

**Figure 3.**
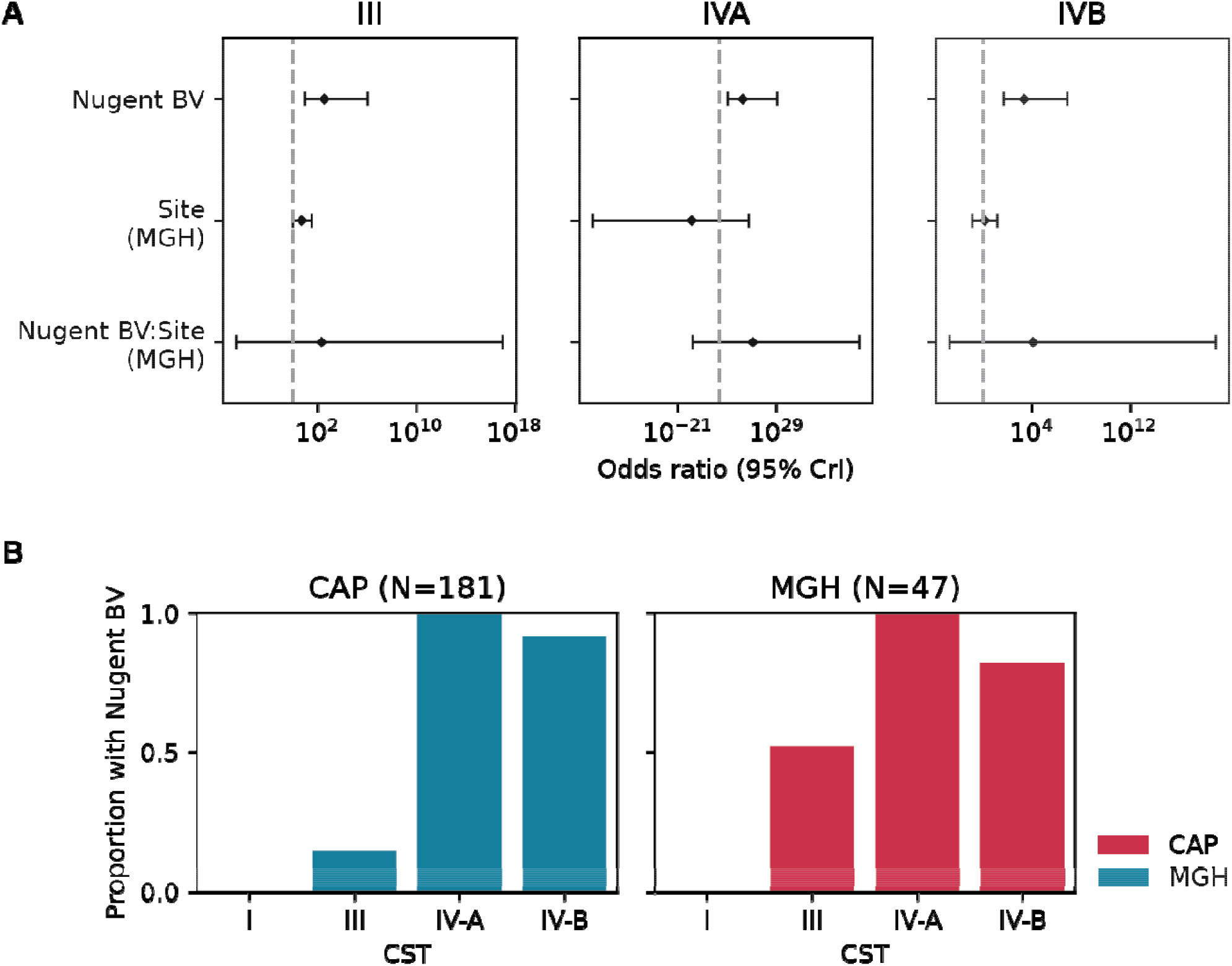
Nugent BV status associates CST without site-specific differences. A) Multinomial odds ratios with 95% credible intervals (CrIs) shown for each predicted CST with CST I as the baseline. Fixed effects included Nugent BV status, the interaction of Nugent BV status and site, and site (with CAPRISA as the baseline). Participant ID was included as a random effect. All screening and follow up visit samples with paired 16s rRNA gene sequencing data were included in this analysis (N=228 total samples; 181 samples from CAP and 47 samples from MGH). B) Proportion of Nugent BV across CST groups for CAPRISA (left) and MGH (right).

To explore potential drivers of the site-specific differences in Amsel diagnosis, we examined each individual Amsel component – pH, clue cells, characteristic discharge, and whiff test – across CSTs and Nugent categories (normal 0-3, intermediate 4-6, BV 7-10) by site (Figure 4). Among samples with Nugent BV, each of the four Amsel criteria were less frequently positive at CAPRISA than at MGH. When stratified by CST rather than Nugent category, the proportion of reported discharge was lower at CAPRISA across CST IV-A and CST IV-B. We also observed a slightly lower proportion of clue cells in CST IV-B samples at CAPRISA, despite this CST being characterized by high *Gardnerella* abundance and clue cells often being attributed to epithelial cells studded with *Gardnerella* bacteria. Similarly, elevated vaginal pH and a positive whiff test were likewise less frequently observed in CST IV-A at CAPRISA than at MGH, and elevated pH was also less frequent in CST IV-B at CAPRISA. Together, these patterns point to systematic differences in the presence and/or assessment of individual Amsel components between sites and contribute to the limited reproducibility of Amsel criteria for detecting CST-defined VD across settings. In contrast, Nugent scoring more consistently captured CST-defined VD and was not modified by site.

**Figure 4.**
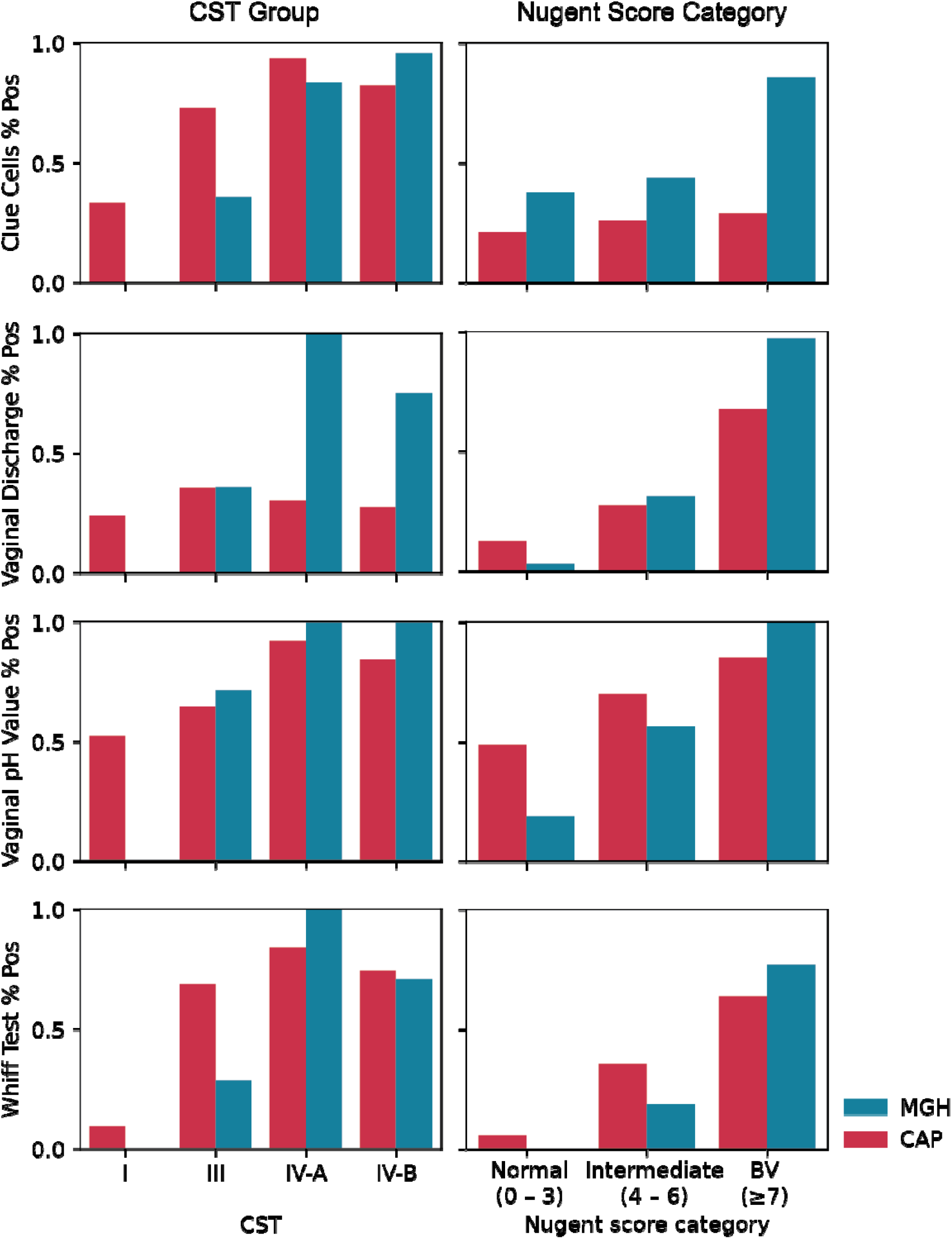
Differences in individual Amsel criteria positivity across CST groups and Nugent score categories. For CST groups, all screening and follow-up visit samples with paired 16s rRNA gene sequencing data were included in this analysis (N=228 total samples; 181 samples from CAP and 47 samples from MGH). For Nugent score categories, all screening and follow up visit samples were included in this analysis (N=557 total samples; 474 samples from CAP and 83 samples from MGH).

## Discussion

Advances in molecular sequencing of the vaginal microbiome has shifted the focus of research from the study of clinical BV to identifying microbial communities associated with increased risk for adverse reproductive health outcomes. As microbiome-directed interventions gain momentum, it is important to understand how clinical diagnostic criteria, traditional microscopic scoring systems, and sequencing-based community classifications relate to one another in appropriately identifying target patient populations. We compared the ability of Amsel criteria, an established diagnostic framework for clinical BV, and Nugent score, a commonly used research criterion for BV, to detect CST-defined VD in two geographically distinct populations. Our analyses yield several key observations: 1) Nugent-based BV more faithfully tracked CSTs than Amsel criteria; 2) CSTs did not clearly distinguish Amsel positivity within Nugent BV positive communities; and 3) the association between Amsel criteria and CST differed by site. Collectively, these findings emphasize that the choice of diagnostic framework should be aligned with the goals of an intervention: while Amsel criteria may be appropriate for diagnosing symptomatic BV, they may be less suitable for detecting asymptomatic BV and vaginal dysbiosis (VD) or at-risk microbiome states across diverse settings and geographies.

Epidemiologic studies linking vaginal microbiota to adverse outcomes (such as preterm birth, infertility, and HIV acquisition) have frequently relied on Nugent scoring and, more recently, sequencing-based CSTs to define microbiome community states^11-15^. Our data show that Nugent scoring aligns more consistently than Amsel criteria with sequencing-defined community structure, particularly in distinguishing *Lactobacillus*-dominant from non-*Lactobacillus*-dominant communities. This finding is consistent with prior reports that Nugent scoring is more sensitive than Amsel criteria for detecting diverse, non-*Lactobacillus*-dominant communities^25,29-34^. Within this context, the discordant diagnostic groups (A+N- and A-N+) further highlight limitations of both systems. A+N-samples, which were predominantly CST III, suggest that clinical signs captured by Amsel criteria can arise in the context of *L. iners*-rich communities which will appear normal or intermediate by Nugent score. Conversely, A-N+ samples exhibited microbiota compositions similar to A+N+ samples, underscoring the lower sensitivity of Amsel criteria for detecting VD-associated communities.

We also observed pronounced site-specific differences in how Amsel and Nugent criteria related to sequencing-defined CSTs. Amsel criteria tracked CST-defined VD reasonably well at MGH but substantially less at CAPRISA, where many CST IV communities lacked Amsel positivity. By contrast, Nugent score showed robust associations with CST III, CST IV-A, and CST IV-B at both sites. These patterns did not appear to be explained by differences in CST distributions with the higher proportion of CST IV-A at CAPRISA. Instead, they appeared to be driven by a difference in the positivity of individual Amsel components among participants with CST IV between sites, most commonly less discharge at CAPRISA. These disparities may reflect differences in local biological and behavioral factors that modulate clinical signs despite similar microbiome compositions. Potential contributors include variation in hormonal contraceptive use, host immune responses, or environmental exposures.

These findings have direct implications for the evaluation of microbiome-targeted therapies^11-15^. Current regulatory guidance for BV treatment trials emphasizes Amsel criteria for defining clinical BV and recurrent disease^35^, and there is no regulatory body that recognizes VD as an established indication or treatable risk-factor for disease. Our results indicate that a substantial proportion of women with CST-defined VD would be missed by Amsel-based diagnosis, particularly in settings where VD is common. Thus, interventions aimed at correcting VD to reduce preterm birth, STI acquisition, or recurrent BV should identify their target populations using Nugent score and/or sequencing-based definitions rather than Amsel criteria. As we gain a greater understanding of the role of *L. iners*, the ability of sequencing methods to distinguish this from other lactobacilli may drive the use of molecular methods to identify VD. Analogous to the measurement and treatment of elevated cholesterol levels to prevent cardiovascular events, we anticipate that future strategies will increasingly focus on identifying and modifying at-risk microbiome states to improve reproductive health.

A major strength of this study is the integration of clinical, microscopic, and sequencing-based measures across two geographically and demographically distinct cohorts. This design allowed us to compare diagnostic frameworks using a common reference (CSTs) and to directly assess site-specific differences. Limitations include the reliance on relative abundance sequencing data, which cannot distinguish changes in total bacterial load or changes in the absolute abundance of key taxa, and therefore, may obscure the contribution of absolute bacterial burden to clinical signs. Furthermore, while our findings highlight important patterns, the trial was not originally powered to identify all potential sources of site-level heterogeneity, and we cannot fully disentangle biological from operational contributors to variability in Amsel criteria assessment or Nugent scoring.

In summary, our results delineate important differences between Amsel and Nugent criteria in their alignment with sequencing-defined vaginal microbial communities and demonstrate that Amsel performance varies by population. For efforts aimed at improving reproductive health through microbiome-directed interventions, it will be essential to move beyond symptom-based definitions of BV toward microbial community-based frameworks that sensitively detect at-risk vaginal microbiome communities regardless of clinical presentation. Such an approach will be critical for designing inclusive, globally relevant interventions and for accurately assessing their impact on women’s health.

## Methods

### Study design and participants

This was an exploratory analysis conducted on samples collected from a randomized clinical trial (VIBRANT)^12^. Briefly, the trial enrolled participants at two sites: CAPRISA Research Clinic in Vulindlela, South Africa, and Massachusetts General Hospital in Boston, US.

Potential participants were invited to the clinic for a screening visit (South Africa) or pre-screened over the phone prior to presenting for an in-person screening visit (Boston). Participants who provided informed consent then self-collected vaginal swabs for Amsel criteria and Nugent scoring, and partway through the study an additional swab was collected and placed in a nucleic acid stabilizing buffer (C2 buffer)^28^. One swab was placed in a vial with saline, which was used to score Amsel criteria, while the second swab was used to prepare the vaginal smear for Nugent scoring and pH assessment using a pH indicator strip. The study was reviewed and approved by the South African Health Products Regulatory Authority (20230615) and the Food and Drug Administration (IND 029629), as well as local ethics committees, University of KwaZulu-Natal’s Biomedical Research Ethics Committee and the Mass General Brigham Institutional Review Board (BREC/00005620/2023; MGB IRB 2023P001035).

### BV Diagnosis by Amsel criteria and Nugent scoring

At CAPRISA, vaginal swabs suspended in saline were transported to the site laboratory, where a drop of the sample was placed onto a glass slide, cover slipped and examined microscopically for clue cells. At the Boston site, this procedure was performed on-site by the study clinician. At both sites, 10% potassium hydroxide (KOH) solution was added to a separate aliquot of vaginal fluid to assess the presence of a fishy odor (indicating a positive whiff test). Both sites received standardized training, and inter-site performance was periodically evaluated to ensure procedural consistency and quality assurance. BV by Amsel’s criteria was defined as being positive for at least three or more criteria.

Vaginal smears on glass slides were air-dried, heat-fixed and Gram-stained following the standard protocol, then examined under 100× oil immersion microscopy, and bacterial morphotypes were quantified based on the Nugent criteria^22^. Each slide was independently evaluated by at least two readers. Discrepancies were resolved by a third reader. Teams at both sites were trained by the same PI, and cross-site blinded evaluations were performed to assess scoring concordance. BV by Nugent score was defined as 7–10, intermediate as 4–6, and normal as 0–3.

### DNA extraction

Vaginal swabs in C2 buffer underwent automated DNA extraction using the MagAttract PowerMicrobiome DNA/RNA EP Kit (Qiagen, Germantown, MD, USA) automated on a Microlab STAR robotic platform (Hamilton, Reno, NV, USA) as described previously^12^. Each run included negative controls (nuclease-free water and unused swabs in C2 buffer) and positive controls consisting of two defined mock communities: Mock1 (15 *L. crispatus* strains plus a standardized vaginal microbial community) and Mock2 (15 *L. crispatus* and selected bacterial isolates). DNA was purified via magnetic bead separation, eluted in nuclease-free water and quantified using a Qubit fluorometer (Thermo Fisher Scientific, USA) to confirm sufficient yield and quality for 16S rRNA gene sequencing.

### 16S rRNA gene amplicon sequencing and taxonomy assignment

16S rRNA gene amplicon libraries were prepared following the Illumina 16S Metagenomic Library preparation protocol, with all samples processed alongside positive (ZymoBIOMICS™ Microbial Community Standard) and negative (nuclease-free water) controls. The V3-V4 hypervariable region of the bacterial 16S rRNA gene was amplified in a two-step PCR using 319F/806R primers containing Illumina overhang adapters and unique dual indices for multiplexing. Amplicons (∼627 bp) were visualized by agarose gel electrophoresis, pooled based on band intensity, and purified using Agencourt AMPure XP magnetic beads Purification Kit (Beckman Coulter, Brea, CA,USA). Library integrity and fragment size distribution were assessed using the Agilent TapeStation system. Taxonomic assignment was performed from demultiplexed reads using standard bioinformatic pipelines for 16S rRNA gene amplicon analysis^36^. The Genome taxonomy database (GTDB_bac120_arc53_ssu_r207_fullTaxo.fa)^37^ was used for taxonomic assignment up to the species level. The resulting ASV table, taxonomic table and relevant metadata were consolidated into a phyloseq object using the phyloseq R package^38^. Community state types were inferred using VALENCIA (VAginaL community state typE Nearest CentroId clAssifier)^39^, implemented in python (version 3.6).

#### Statistical analysis

Data analyses and visualization were performed using pandas 2.2.2, numpy 1.26.4, matplotlib 3.9.2, seaborn 0.13.2, skbio 0.6.3, and sklearn 1.7.0 in Python 3.12.3, as well as dplyr 1.1.4, tibble 3.3.0, stringr 1.5.1, forcats 1.0.0, ggplot2 3.5.2, ggh4x 0.3.1, and scales in R 4.5.0. Bayesian multinomial mixed-effects logistic regression models were fit in R using the brms package (v2.23.0), with community state type (CST) as the outcome and Nugent (or Amsel) BV status, site, and their interaction as fixed effects, and a random intercept for participant: CST ∼ Nugent (or Amsel) BV status + site + Nugent (or Amsel) BV status × site + (1 | participant). CST I and CAPRISA were used as the reference levels.

## Data Availability

Source data and original code is deposited at Zenodo and publicly available and accessible under 10.5281/zenodo.18111510

## Data and Code Availability

Source data and original code will be deposited at Zenodo and publicly available and accessible under 10.5281/zenodo.18111510.

## Funding

This project was funded by the Gates Foundation (INV-019055 and INV-037901 to CM, and INV-037902 to DP).

## Acknowledgments

We express deep appreciation for the tangible and intangible contributions of all members of the Vaginal Microbiome Research Consortium who have supported this work over many years with encouragement, suggestions, and critiques, and specifically the contributions of Dr. Emily Balskus, Dr. Daniel Erchick, Dr. Indriati Hood-Pishchany, Dr. Margaret Kasaro, Dr. Moses Obimbo, Dr. Seth Rakoff-Nahoum, Dr. David Relman, Dr. Katharina Ribbeck, and Dr. Sean Stowell.

## Author contributions

Conceptualization: DP, SN, CMM

Developed the study protocol: DP, LS, SN, LL, CC, JAP, AUH, NoM, DSK, CMM

Conducted the clinical trial: DP, CC, NoM, MM, AK, BCD, CMM

Processed and evaluated trial samples: SN, AM, AK, NzM, NiM, GM, MM, BCD

Generated laboratory data for trial samples: SN, AMP, AM, AK, NzM, NiM, GM, AK, BCD, JX

Performed analysis: MZ, JE, JS, AK, LV

Writing of initial draft: MZ, CMM

Reviewing of the final manuscript: all authors

## Competing Interests

CMM has been a consultant for DIVA Inc, Freya Biosciences, Novonesis and is on the scientific advisory boards of Concerto Biosciences and Ancilia Biosciences. JAP holds a patent for a device to diagnose genital inflammation. The other authors have no competing interests.

## Supplemental Text Figures

**SFigure 1.**
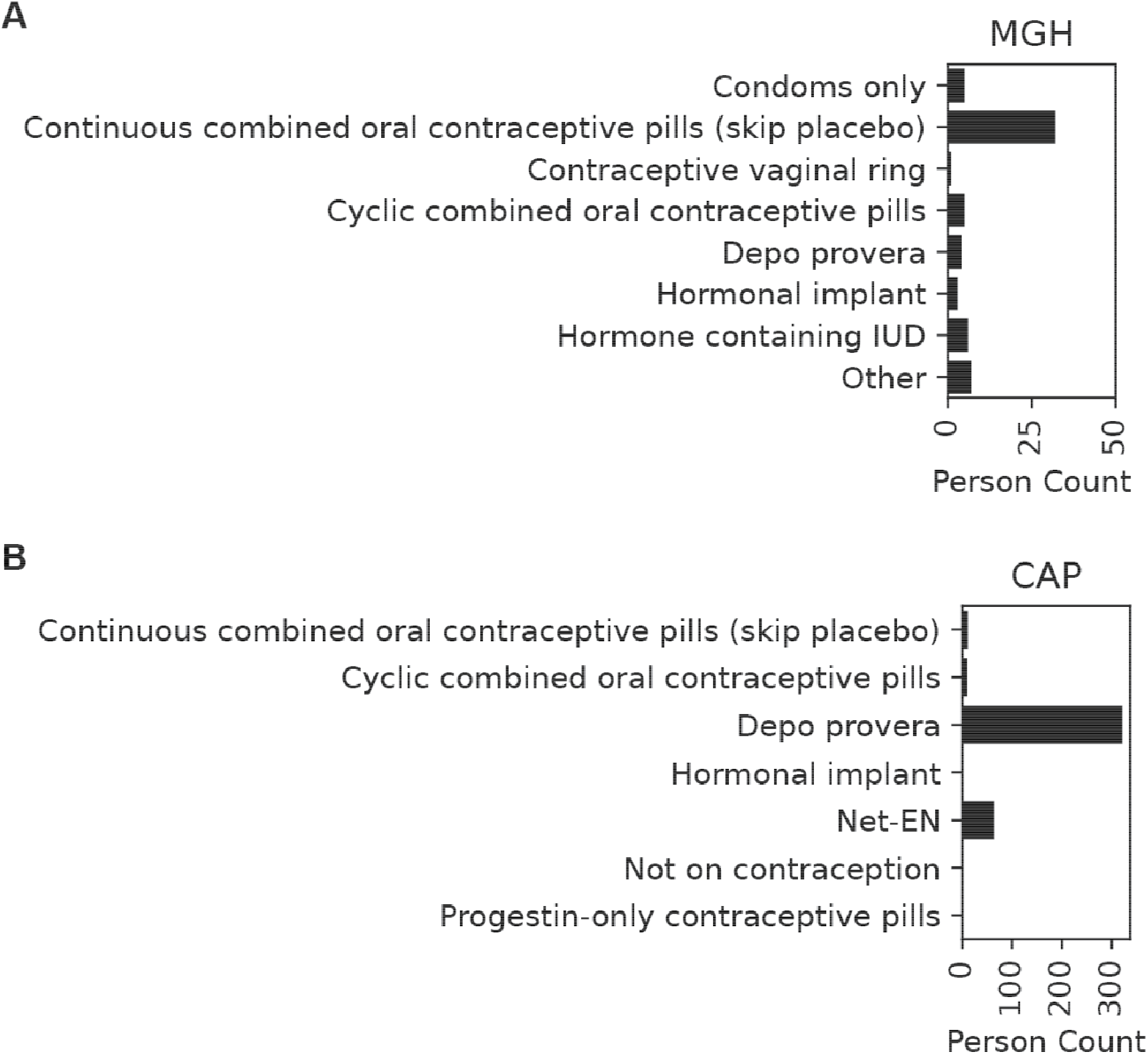
Contraception use of sample donors across sites. A) CAPRISA B) MGH

**SFigure 2.**
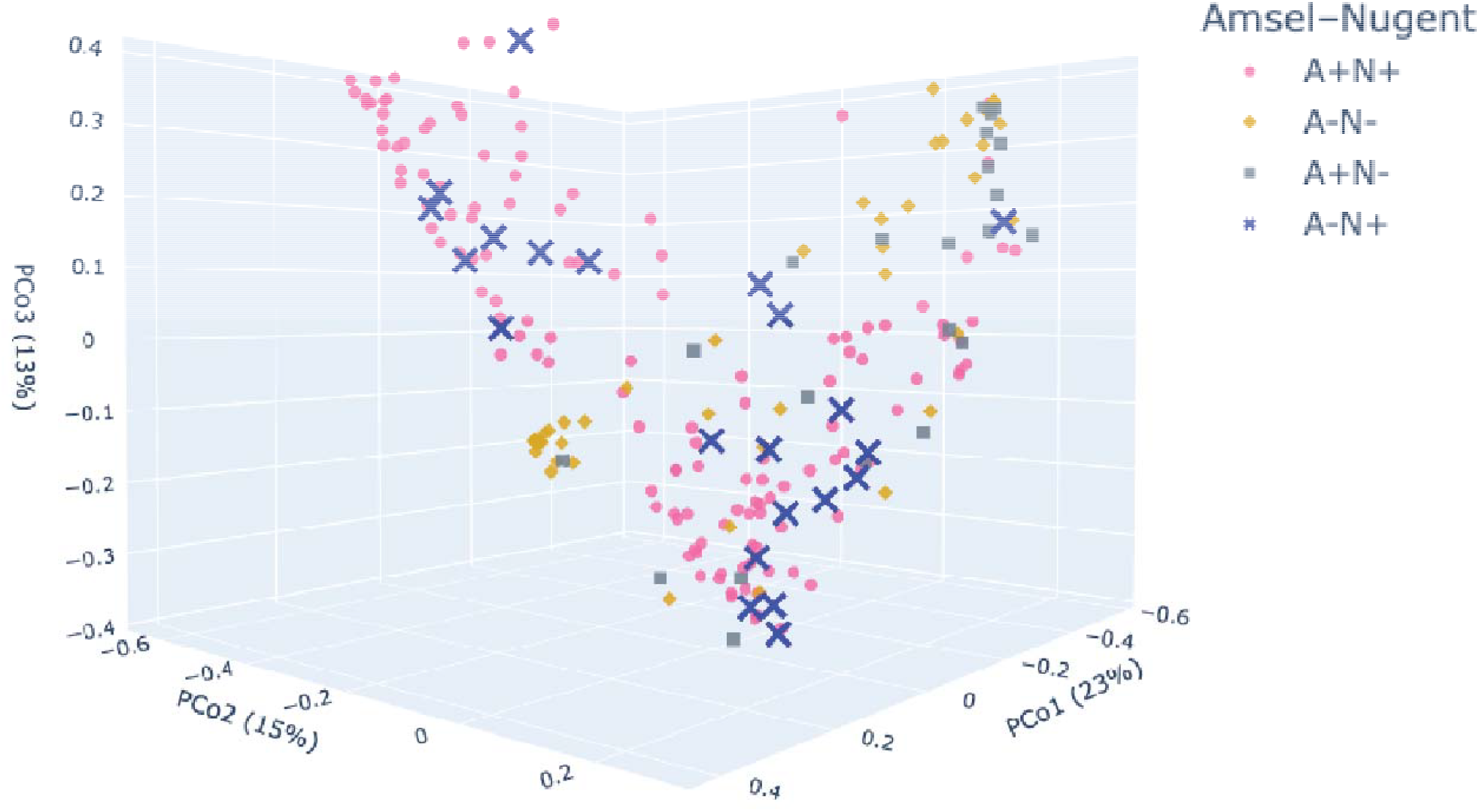
3D PCoA colored by Amsel/Nugent Status.

